# Sodium-glucose cotransporter 2-associated perioperative ketoacidosis: a protocol for SAPKA systematic review

**DOI:** 10.1101/2022.05.22.22275348

**Authors:** Hiroyuki Seki, Satoshi Ideno, Toshiya Shiga, Hidenobu Watanabe, Motoaki Ono, Akira Motoyasu, Hikari Noguchi, Kazuya Kondo, Takahiro Yoshikawa, Hiroshi Hoshijima, Shunsuke Hyuga, Miho Shishii, Ai Nagai, Midoriko Higashi, Takashi Ouchi, Kazuki Yasuda, Norifumi Kuratani

## Abstract

**Background:** In recent years, reports have increased regarding patients treated with sodium-glucose cotransporter 2 inhibitors (SGLT2i) who develop severe ketoacidosis during the perioperative period. This systematic review aims to extrapolate precipitating factors from case reports of SGLT2i-associated perioperative ketoacidosis.

**Methods:** Two independent researchers will search PubMed, EMBASE, and Web of Science, up to June 1, 2022, with no language restrictions, to identify reports of perioperative ketoacidosis associated with SGLT2i use. We will include case reports/series describing patients taking SGLT2i who developed ketoacidosis (defined as blood pH < 7.3 and blood or urine ketone positivity) pre-, intra-, or postoperatively up to 30 days following surgery. Patients for whom SGLT2i was newly prescribed postoperatively and those for whom the diagnosis of ketoacidosis does not fulfill our criteria will be excluded.

**Competing interest statement:** Dr. Yasuda reports receiving grants from Ono Pharmaceutical and Mitsubishi Tanabe Pharma Corporation.

**Funding statement:** The Japan Society for the Promotion of Science Grant-in-Aid for Scientific Research (C) (Grant Number 21K06676) funded this study.

## Introduction

Sodium-glucose cotransporter 2 inhibitors (SGLT2i) comprise an anti-diabetic drug class. Aside from lowering the blood glucose levels, this type of drug also reduces the hemoglobin A1c level, promotes weight loss, lowers blood pressure, and provides cardiorenal protection^1)^. However, these inhibitors have been associated with adverse events such as ketoacidosis, which is fatal. Despite the increasing number of reports on SGLT2i-associated ketoacidosis, specifically during the perioperative period, its etiology and risk factors have not been fully elucidated. As the prescription of SGLT2i becomes more ubiquitous, it is necessary to understand the clinical features of SGLT2i-associated ketoacidosis. This systematic review aimed to extrapolate the precipitating factors for SGLT2i-associated perioperative ketoacidosis (SAPKA) by summarizing the currently available case reports.

## Methods

This study will be conducted in compliance with the preferred reporting items systematic reviews and meta-analysis 2020 statement^2)^. The preliminary search has been completed, and the formal search will start on June 1, 2022. The anticipated date of completion is December 31, 2022.

### Eligibility criteria

We will include case reports/series involving patients taking SGLT2i who developed ketoacidosis (defined as a blood pH <7.3 and blood or urine ketone positivity) preoperatively, intraoperatively, or up to 30 days postoperatively. Letters, reviews, and conference abstracts will be included. Patients newly prescribed with SGLT2i after surgery and those who do not fulfill the criteria for ketoacidosis will be excluded. Cases in which pH value is unclear, but the authors state “acidosis” will be included.

### Search strategy and study selection

Studies will be obtained from electronic databases, including PubMed, EMBASE, and Web of Science, by two independent researchers (H.S. and S.I.). The search will be conducted on June 1, 2022, without language restrictions for identifying perioperative ketoacidosis associated with SGLT2i. The full search strategy is described in Table 1. The reference lists of all identified studies and those of previous meta-analyses on similar topics will be checked. The titles and abstracts of the obtained references will be independently screened by two reviewers (H.S. and S.I.), and the full-text articles of relevant abstracts will be collected. Disagreements will be resolved by discussion or consultation with a third author (K.Y.).

**Table 1.** Search strategies for each database

|  |  |
| --- | --- |
| PubMed | ("SGLT2 inhibitors"[All Fields] OR "sodium-glucose cotransporter 2 inhibitors"[All Fields] OR ("canagliflozin"[MeSH Terms] OR "canagliflozin"[All Fields]) OR ("dapagliflozin"[Supplementary Concept] OR "dapagliflozin"[All Fields] OR "dapagliflozin s"[All Fields]) OR ("empagliflozin"[Supplementary Concept] OR "empagliflozin"[All Fields]) OR ("ertugliflozin"[Supplementary Concept] OR "ertugliflozin"[All Fields]) OR ("ipragliflozin"[Supplementary Concept] OR "ipragliflozin"[All Fields]) OR ("1 5 anhydro 1 5 4 ethoxybenzyl 2 methoxy 4 methylphenyl 1 thioglucitol"[Supplementary Concept] OR "1 5 anhydro 1 5 4 ethoxybenzyl 2 methoxy 4 methylphenyl 1 thioglucitol"[All Fields] OR "luseogliflozin"[All Fields]) OR ("2s 3r 4r 5s 6r 2 4 chloro 3 4 ethoxybenzyl phenyl 6 methylthio tetrahydro 2h pyran 3 4 5 triol"[Supplementary Concept] OR "2s 3r 4r 5s 6r 2 4 chloro 3 4 ethoxybenzyl phenyl 6 methylthio tetrahydro 2h pyran 3 4 5 triol"[All Fields] OR "sotagliflozin"[All Fields]) OR ("6 4 ethylphenyl methyl 3 4 5 6 tetrahydro 6 hydroxymethyl spiro isobenzofuran 1 3h 2 2h pyran 3 4 5 triol"[Supplementary Concept] OR "6 4 ethylphenyl methyl 3 4 5 6 tetrahydro 6 hydroxymethyl spiro isobenzofuran 1 3h 2 2h pyran 3 4 5 triol"[All Fields] OR "tofogliflozin"[All Fields])) AND ("ketosis"[MeSH Terms] OR "ketosis"[All Fields] OR "ketoacidosis"[All Fields] OR ("acidosis"[MeSH Terms] OR "acidosis"[All Fields] OR "acidoses"[All Fields])) |
| Web of Science | ("SGLT2 inhibitors" OR "sodium-glucose cotransporter 2 inhibitors" OR canagliflozin OR dapagliflozin OR empagliflozin OR ertugliflozin OR ipragliflozin OR luseogliflozin OR sotagliflozin OR tofogliflozin) AND (ketoacidosis OR acidosis) |
| EMBASE | ("SGLT2 inhibitors" OR "sodium-glucose cotransporter 2 inhibitors" OR canagliflozin OR dapagliflozin OR empagliflozin OR ertugliflozin OR ipragliflozin OR luseogliflozin OR sotagliflozin OR tofogliflozin) AND (ketoacidosis OR acidosis) |

### Data extraction

Patient data, including age, sex, body mass index, purpose of SGLT2i prescription, comorbidities, type of diabetes, type of SGLT2i, anti-diabetic and other medications, type of surgery and anesthesia, data on withholding the agents before surgery, nature of presentation as ketoacidosis, biochemical parameters (including serum glucose, hemoglobin A1c, pH, serum bicarbonate, partial pressure of arterial carbon dioxide, anion gap, plasma and urine ketones, and glycosuria), details of management, complications, and outcome, will be independently extracted by 14 reviewers (H.S., S.I., H.W., M.O., A.M., H.N., K.K., T.Y., H.H., S.H., M.S., A.I., M.H., and T.O.) working in seven teams. In this study, ketoacidosis is defined as a blood pH <7.30, along with blood or urine ketone positivity based on several guidelines^3)^. Disagreements will be resolved by discussion or consultation with a third author (K.Y.).

### Assessment of methodological quality of the cases

The methodological quality of the cases will be assessed using a previously published method^4)^. The quality of the report will be graded from zero (low) to five (high). Disagreements will be resolved by discussion or consultation with a third author (K.Y.).

### Strategy for data synthesis

A narrative synthesis of the findings of the included studies will be provided. It will be structured around the type of intervention, target population characteristics, type of outcome, and intervention content.

## Data Availability

All data produced in the present study will be available upon reasonable request to the authors.

## Dissemination plans

The findings of this study will be submitted to peer-reviewed journals, and abstracts will be submitted to relevant national and international meetings.

## Author’s contribution

Review design/planning: H.S., S.I., N.K., T.S., K.Y.

Data extraction/analysis: H.S., S.I., H.W., M.O., A.M., H.N., K.K., T.Y., H.H., S.H., M.S., A.I., M.H., T.O.

Writing manuscript: H.S., N.K., T.S., K.Y. Critical review: All authors

## Acknowledgement

None.

## Declaration of interest

Dr. Yasuda reports grants from Ono Pharmaceutical and Mitsubishi Tanabe Pharma Corporation.

## Funding

This work was supported by the Japan Society for the Promotion of Science Grant-in-Aid for Scientific Research (C) (Grant Number 21K06676).

## Notes

### Author Declarations

The study will use ONLY openly available human data that were originally published as research papers.

